# BrcaDx: Precise identification of breast cancer from expression data using a minimal set of features

**DOI:** 10.1101/2023.03.04.23286744

**Authors:** Sangeetha Muthamilselvan, Ashok Palaniappan

## Abstract

**Background:** Breast cancer is the foremost cancer in worldwide incidence, surpassing lung cancer notwithstanding the gender bias. One in four cancer cases among women are attributable to cancers of the breast, which are also the leading cause of death in women. Reliable options for early detection of breast cancer are needed.

**Methods:** Using public-domain datasets, we screened transcriptomic profiles of breast cancer samples, and identified progression-significant linear and ordinal model genes using stage-informed models. We then applied a sequence of machine learning techniques, namely feature selection, principal components analysis, and k-means clustering, to train a learner to discriminate ‘cancer’ from ‘normal’ based on expression levels of identified biomarkers.

**Results:** Our computational pipeline yielded an optimal set of nine biomarker features for training the learner, namely NEK2, PKMYT1, MMP11, CPA1, COL10A1, HSD17B13, CA4, MYOC, and LYVE1. Validation of the learned model on an internal testset yielded a performance of 99.5% accuracy. Blind validation on an external dataset yielded a balanced accuracy of 95.5%, demonstrating that the model has effectively reduced the dimensionality of the problem, and learnt the solution. The model was rebuilt using the full dataset, and then deployed as a web app for non-profit purposes at: https://apalania.shinyapps.io/brcadx/. To our knowledge, this is the best-performing freely available tool for the high-confidence diagnosis of breast cancer, and represents a promising aid to medical diagnosis.

## Introduction

Breast cancer is the most commonly diagnosed cancer in the world, with a staggering 2.3 million cases in 2020^1^. It accounts for approximately 24.5% of cancer cases and 15.5% of cancer deaths among women, ranking #1 in both incidence and mortality in most countries. Modelling studies predict an exponential and asymmetric rate of increase in breast cancer incidence among low human development index (HDI) nations relative to high HDI nations, due to an unmitigated increase in risk factors in low HDI nations^2^. In India, for e.g., the age of onset of breast cancer has advanced ten years earlier relative to that in Europe and America. About 29% - 52% of women with breast cancer in India present in the more severe advanced stages, leading to poor prognosis^3^. Low HDI nations are likely to also suffer from problems due to the lack of social awareness and existent taboos, especially in rural areas. Alternative diagnostic methods based on a minimal set of biomarkers are urgently needed to effectively redress the situation^4^.

The advent of –omics data has ushered in AI-based approaches to cancer diagnosis. However, contemporary AI-based diagnostic methods are saddled with unreasonable dimensionality of the hypothesis space, and typically require sequencing of hundreds of biomarkers to achieve clinical utility. Dimensionality reduction techniques like principal components (PC) analysis are generally used for extracting optimal feature subsets, especially when linear relationships exist in the dataset. PC analysis has been earlier used to detect multiple cancer types simultaneously, with a costly compromise in accuracy and interpretation^5^.

Working in the space of PCs tends to lead to more robust clustering outcomes^6^, and k-means clustering is an effective technique for analyzing transformed spaces^7,8^. Building on the above observations, this study has two principal objectives: (i) develop and validate the most efficient integrative computational pipeline for breast cancer classification based on a minimal hypothesis space; and (ii) translate the resulting diagnostic classifier into a web-app service to aid medical decision-making.

## Materials and Methods

The overall workflow is summarised in Fig. 1 and discussed in detail below.

**Fig 1.**
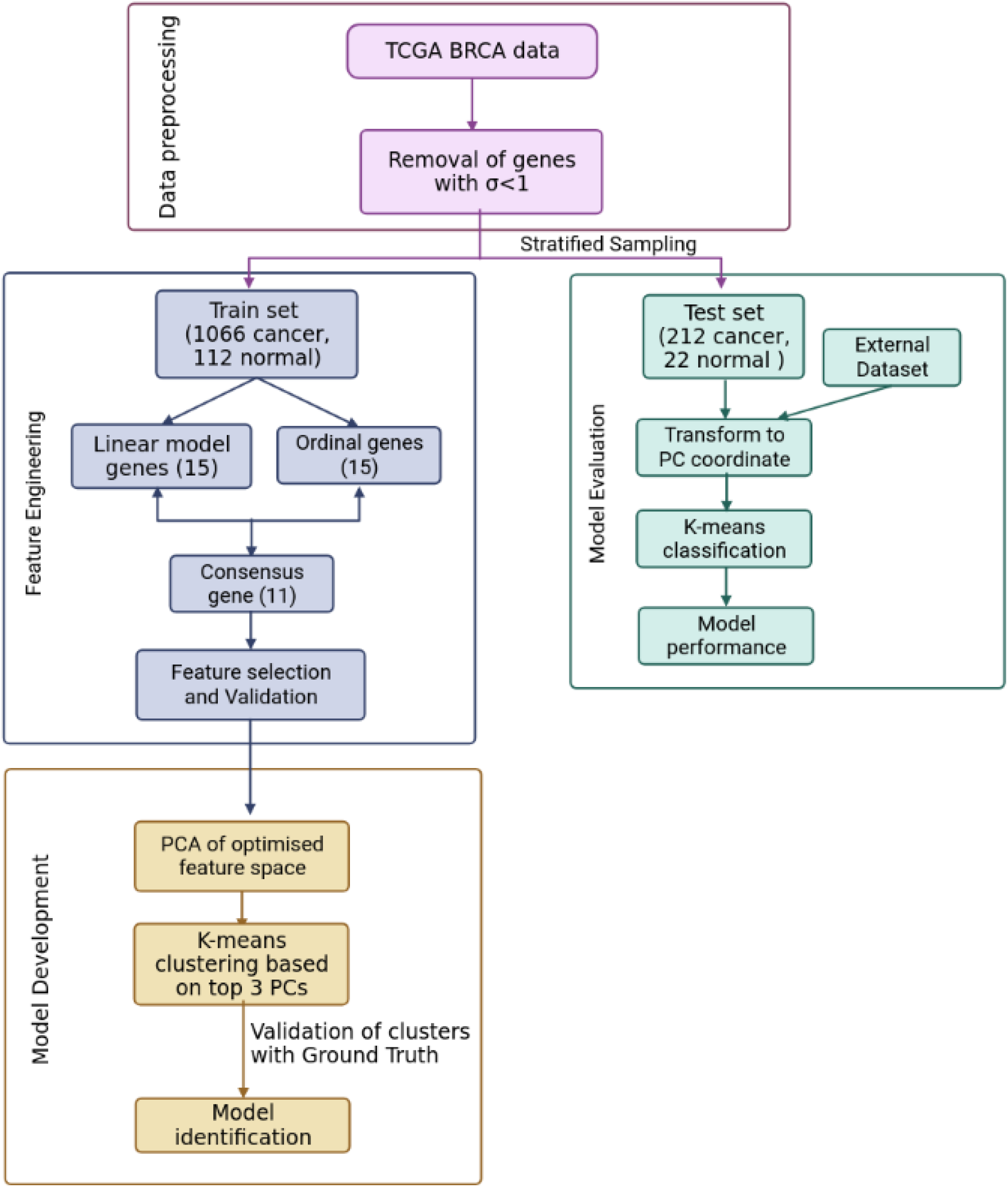
ML pipeline used in the study for the design of a simple, effective and optimal cancer vs normal classifier.

### Data Pre-processing

RSEM-normalised BRCA expression dataset (gdac.broadinstitute.org_BRCA.Merge_rnaseqv2_illuminahiseq_rnaseqv2_unc_edu_Level_3 RSEM_genes_normalized_data.Level_3.2016012800.0.0.tar.gz) was retrieved from the TCGA using firebrowse portal^9^ by selecting the Cohort as ‘Breast invasive carcinoma’. The samples were annotated as ‘normal’ or ‘cancer’ based on the sample-encoding part in the patient barcode (uuid) in the variable ‘Hybridization REF’. The sample stage was extracted from the attribute ‘patient.stage_event.pathologic_stage’ in the associated clinical metadata file retrieved for the same cohort as gdac.broadinstitute.org_BRCA.Merge_Clinical.Level_1.2016012800.0.0.tar.gz. Genes with minimal variation in expression across the samples were removed if the expression σ < 1. The resulting data matrix was then processed through voom in limma to prepare for linear modelling^10^. Then it was split into train: test datasets in the ratio 80:20 stratified on the target class. Data pre-processing was done in R (www.r-project.org).

### Feature Engineering

The training dataset was used to identify the features for the problem. Two models were considered to extract potential features:

(1) A linear model of stagewise expression in each gene was performed using R *limma*^10^, with the following equation

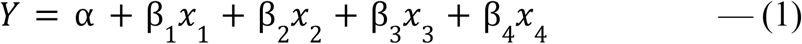

where the intercept α is the baseline expression obtained from the controls, the independent variables are indicator variables of the sample’s stage, and β_i_ are the predicted log fold-change (lfc) coefficients relative to controls. Further the model was subjected to empirical Bayes adjustment for obtaining moderated t-statistics^11^. Multiple hypothesis testing was corrected using the Benjamini Hochberg method^12^.

(2) An ordinal model of gene expression was also considered. Here the cancer stage is treated as a numeric variable according to the equation:

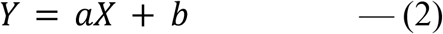

where X is the cancer stage taking the values 0, 1, 2, 3, and 4, corresponding to Control, Stage-1, Stage-2, Stage-3, and Stage-4, respectively.

### Feature space optimization

Genes from the linear and ordinal expression models were ranked based on the adj. p-value. The consensus set between the top-ranked 15 genes of the linear and ordinal models was determined and then subjected to feature selection using Boruta ^13^ and Recursive Feature Elimination^14^ (RFE). Boruta implements a wrapper algorithm based on Random Forest to select features either strongly or weakly connected to the outcome variable, while RFE implements a backward selection process to identify an optimal set of predictors. Post feature-selection, the retained features were validated using variance inflation analysis, involving regressing each independent variable on all the other independent variables in turn, identifying and removing redundancy till a minimal feature space has been obtained ^15^. The variance inflation factor (VIF) score was calculated using:

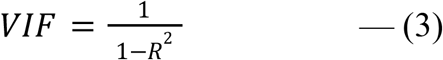

where R^2^ is the goodness-of-fit of the fitted model. A variable with VIF = 1.0 is perfectly independent of all other variables, whereas any variable with VIF > 2.0 was deemed multicollinear with the other variables and iteratively eliminated.

### PCA-based K-Means Clustering

From the validated set of features, the principal components of the subspace spanned by these features were found, and the optimal number of principal components identified using three different criteria, namely scree plot, Kaiser-Guttmann rule^16^, and the proportion of variance explained. K-means clustering with k=2 was performed in the space defined by the optimal principal components, to examine separation between the normal and cancer samples.

### Model evaluation

Classification performance from clustering in the principal components space was evaluated using metrics like accuracy, precision, recall, and F_1_-score. Performance evaluation was done on both the internal testset and an external dataset ‘BRCA-KR’ retrieved from the ICGC DataPortal (https://dcc.icgc.org/). Since BRCA-KR had just three control samples, it was augmented with 218 control samples from GTEx^17^.

## Results

BRCA RNA-Seq data retrieved from TCGA consisted of 1212 samples each with the expression values of 20532 genes. Post data pre-processing, we obtained a dataset of 1178 samples, 18880 genes. We performed an 80:20 stratified sampling of the dataset (with 1066 cancer, 112 normal samples) based on the outcome class to obtain the training dataset (with 854 cancer, 90 normal samples), and test dataset (with 212 cancer, 22 normal samples). The training dataset was voom-processed using limma and then subjected to the two modeling protocols. At an adj.p-value threshold of 1E-5, the linear model yielded 8961 significant genes (Supplementary File S1), while the ordinal model yielded 6888 significant (Supplementary File S2). We examined the overlap among the top 15 genes from each model, which produced eleven consensus genes for subsequent analysis.

Application of the Boruta feature selection protocol on the eleven genes yielded a hypothesis space of only nine genes, while application of the RFE feature selection protocol did not yield any reduction in the size of the hypothesis space. A summary of the final nine consensus genes is presented in Table 1. The hypothesis space was subjected to VIF analysis, to ensure absence of multicollinearity among features, and establish a minimal non-redundant set of features (Table 1, last column). We identified the nine principal components (PCs) of this 9-dimensional space (Table 2), and then visualized the training samples using the top PCs from this analysis (Fig. 2). The application of three PCs clearly resolves and separates the cancer and normal samples (Fig. 2b). To decisively identify the optimal number of PCs, we examined the three criteria outlined in ethods: (i) Kaiser-Guttman criterion yielded top six PCs; (ii) Scree plot showed the first three principal components to be optimal (Fig. 3a); and (iii) the first three PCs explained > 85% variance, passing the proportion of variance explained condition. We reconciled the above findings, and chose the first three principal components to define a 3-dimensional space for applying k-means clustering. Next, we optimized the number of clusters (k) for k-means clustering using the silhouette method^18^ (Fig. 3b). A value of k=2 was obtained, which synchronized with the larger objective to partition the structure of the space into cancer and normal signatures.

**Table 1.** Summary of the consensus features from the two modeling protocols. All features are exceedingly differentially expressed with extreme significance. The largest VIF score does not exceed 1.57, corresponding to a multivariate ‘correlation coefficient’ < 0.6.

| S.No | Feature | lfc | Adj.P.value<br>- linear | Adj.P.value<br>- ordinal | Regulation<br>status | VIF<br>score |
| --- | --- | --- | --- | --- | --- | --- |
| 1 | NEK2 | 4.57 | 2.94E-146 | 6.25E-61 | UP | 1.05 |
| 2 | PKMYT1 | 4.47 | 1.53E-127 | 6.14E-53 | UP | 1.05 |
| 3 | MMP11 | 5.99 | 3.26E-134 | 2.02E-53 | UP | 1.00 |
| 4 | CPA1 | -4.20 | 1.61E-138 | 2.62E-49 | DOWN | 1.54 |
| 5 | COL10A1 | 7.12 | 2.04E-137 | 5.62E-54 | UP | 1.00 |
| 6 | HSD17B13 | -4.86 | 5.67E-117 | 3.71E-51 | DOWN | 1.22 |
| 7 | CA4 | -6.93 | 8.41E-127 | 9.92E-50 | DOWN | 1.57 |
| 8 | MYOC | -6.53 | 3.30E-133 | 4.03E-57 | DOWN | 1.34 |
| 9 | LYVE1 | -4.91 | 3.10E-128 | 3.21E-47 | DOWN | 1.02 |

**Table 2.** Summary of the nine components from the PC analysis, ranked by associated eigenvalue. Cumulative variance enables the application of the ‘proportion of variance explained’ criterion.

| S.No | PC | Eigenvalue | Variance explained (%) | Cumulative variance explained (%) |
| --- | --- | --- | --- | --- |
| 1 | PC1 | 34.487 | 67.24 | 67.24 |
| 2 | PC2 | 7.181 | 14.00 | 81.24 |
| 3 | PC3 | 2.787 | 5.43 | 86.67 |
| 4 | PC4 | 2.039 | 3.97 | 90.65 |
| 5 | PC5 | 1.521 | 2.97 | 93.62 |
| 6 | PC6 | 1.191 | 2.32 | 95.94 |
| 7 | PC7 | 0.887 | 1.73 | 97.67 |
| 8 | PC8 | 0.781 | 1.52 | 99.19 |
| 9 | PC9 | 0.415 | 0.81 | 100 |

**Fig. 2.**
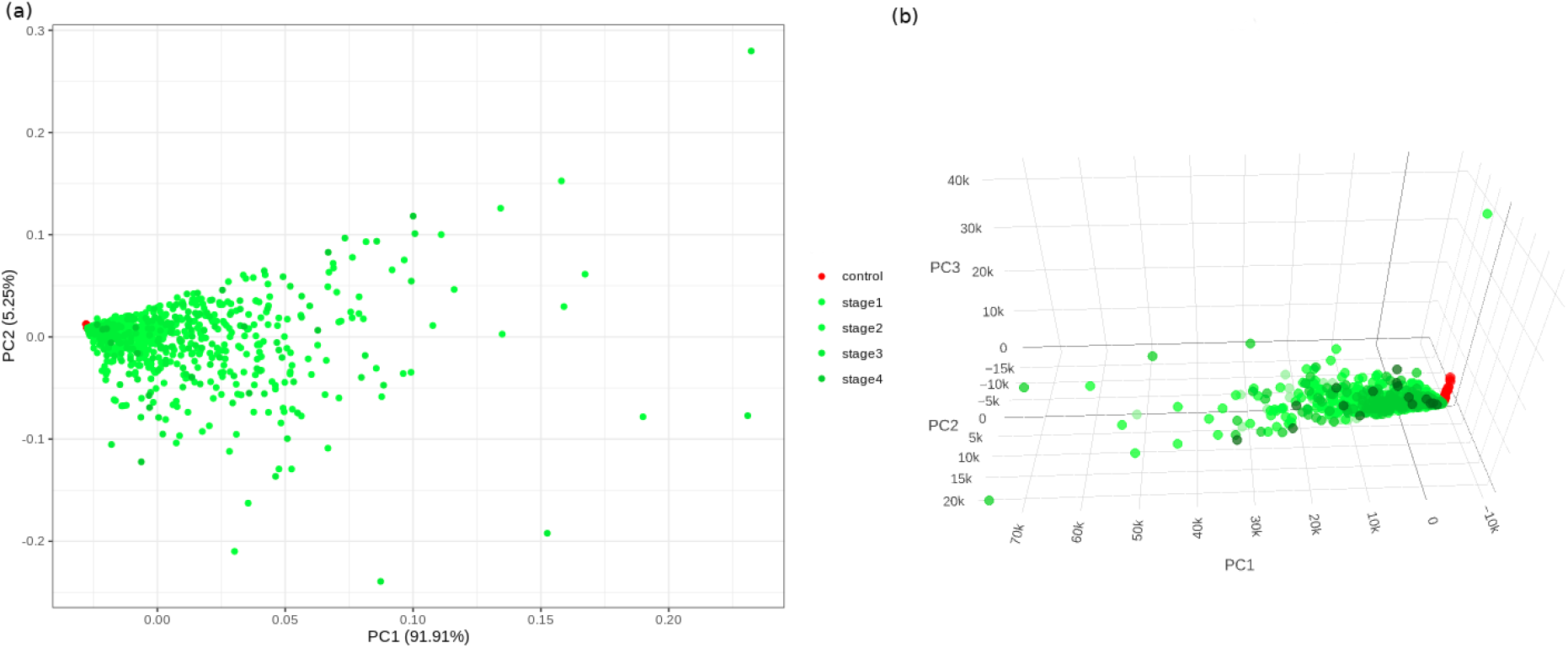
PC analysis of the biomarker expression space. With (a) top two components; and (b) top three components. It is seen that the use of three components expands the separation between the cancer samples and controls in better-defined sub-spaces.

**Fig. 3.**
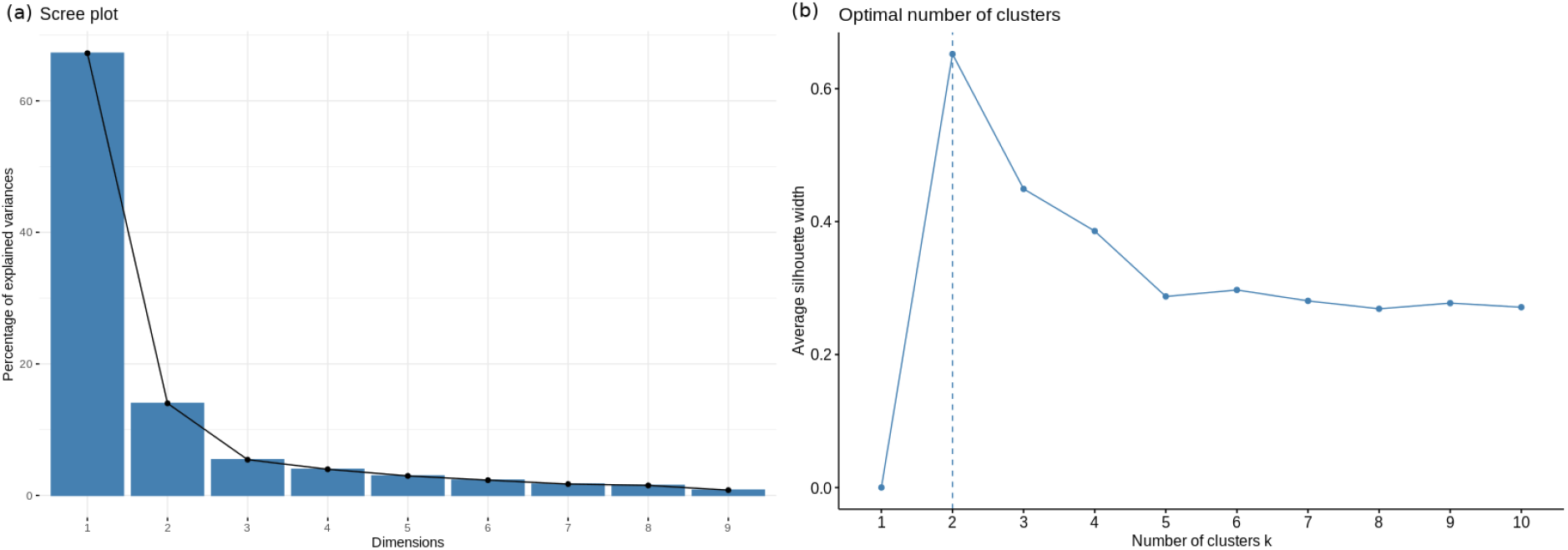
Model parameterization. (a) Scree plot for determination of the optimal number of principal components. The elbow method yields the first three PCs which have a cumulative variance > 85%. (b) Silhouette plot for ascertaining the optimal number of clusters in the structure of the transformed PC-space. The emergent value, k=2, is in sync with the type of problem at hand: binary classification.

From Fig. 4, it is clear that the k-means classifier in the 3-dimensional PC space of the identified biomarkers effectively partitioned the space into cancer vs normal. The performance of the clustering outcomes assessed against the ground truth labels in the training, test and external datasets is presented in Table 2. It is seen that the model produced by the workflow has yielded balanced accuracies of 99.53% and 95.52% on the internal validation and external validation datasets respectively.

**Fig. 4.**
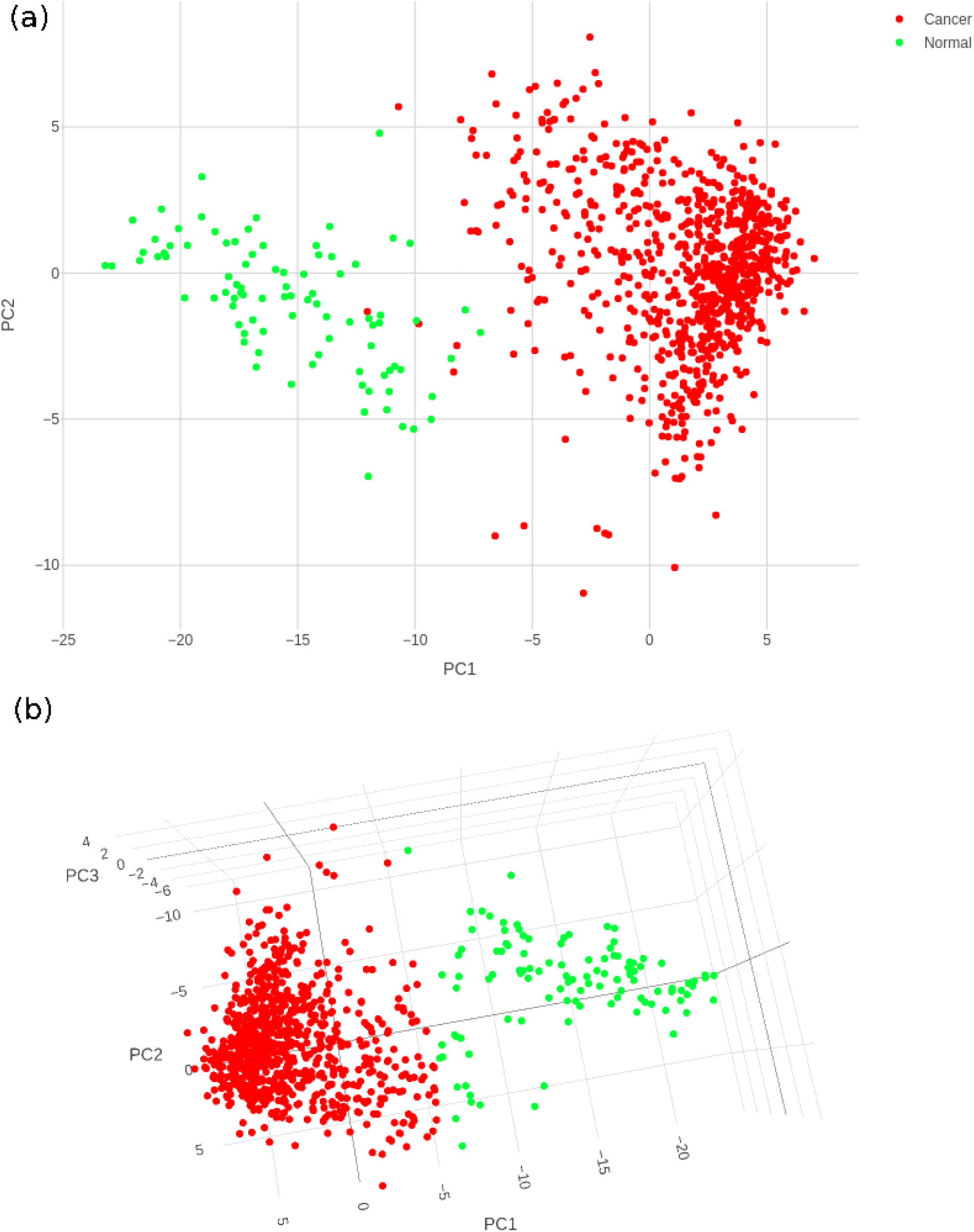
Cancer (red) and control (green) clusters obtained after training the k-means classifier. (a) Two-dimensional projection onto the first two principal components shows some uncertainty in the boundaries of the two clusters; (b) Visualization in the three-dimensional space of the PCs satisfactorily resolves the cluster boundaries.

**Table 2.** Performance metrics of the developed k-means model in the transformed PC space of the identified nine biomarker features. Bal. acc. refers to balanced accuracy, i.e, the average of the accuracies obtained with respect to each class (cancer and normal). F_1_-score is defined as the harmonic mean of the precision and recall. Sensitivity is identical to the recall values.

| S.No | Dataset | Bal. acc. | Specificity | Precision | Recall | $F_1$-score |
| --- | --- | --- | --- | --- | --- | --- |
| 1 | Training | 98.83 | 100 | 100 | 97.66 | 98.81 |
| 2 | Test | 99.53 | 100 | 100 | 99.06 | 99.53 |
| 3 | **External** | 95.52 | 99.55 | 97.73 | 91.49 | 94.51 |

### Deployment

To convert the outcomes in effectively classifying cancer vs normal based on the expression of just a handful of features, we have developed an app, BrcaDx, to freely provide the service to the academic community. BrcaDx is deployed at: https://apalania.shinyapps.io/brcadx/. The model was rebuilt using the full dataset for maximum discriminative performance. Based on an input of the expression of the nine biomarkers, the app carries out a necessary log2 operation of the values, and transforms them into the three-dimensional PC space. The transformed coordinates are fed to the learned k-means clustering model, which locates the sample in either of the two clusters, thus predicting the class of the sample. The app accepts a single-sample input as well as a batch input (samples x biomarkers), in which case it processes all the samples and returns the corresponding prediction for each sample. To facilitate not-for-profit applications, a video tutorial for using the app has been provided on the landing page. The app was implemented using R-Shiny (https://shiny.rstudio.com/).

## Discussion

It is significant to note that some of the biomarkers identified in our study are part of marketed and commercially available signature panels used in the context of breast cancer diagnosis and treatment. Specifically: (i) NEK2 is a constituent of the 11-gene Breast Cancer Index signature used to estimate recurrence^19^; and (ii) MMP11 is a constituent biomarker of the 50-gene Prosigna^20^, and 21-gene OncotypeDX^21^ signature panels, which are both used in estimating likelihood of recurrence. It is interesting to note that the Prosigna panel is based on the PAM50 signature, which is also used to subtype breast cancer into Luminal-A, Luminal-B, HER2-enriched and Basal-like^22^.

The consensus genes used to build our model are known to play key roles in cancers of the breast and other tissues, contributing to breast-cancer specific pathways as well as cancer hallmark processes^23^. The genes NEK2, PKMYT1, and CA4 are known to play indispensable roles in cell cycle progression^24-26^. NEK2 is documented to be overexpressed in breast-cancer tissue relative to normal tissue^27,28^, and is required for the growth, maintenance and survival of the transformed cell^29^. PKMYT1 overexpression is known to be significantly correlated with BRCA subtypes, and indicative of poor prognosis^30^. Downregulation of CA4 is associated with poor prognosis in cancers other than that of the breast, notably uveal melanoma, renal cell cancer, glioma, and lung adenocarcinoma^30,31^, hinting its role in hallmark processes common to many cancers, and its potential significance in establishing such hallmarks in breast cancer progression. Hypermethylation of the CPA1 gene in breast cancer cells has been earlier demonstrated^32,33^, which could lead to its significant downregulation noted here. Recently, COL10A1 was identified as an overexpressed predictive biomarker for breast cancer coexpressed with LRRC15^34^. COL10A1 protein is a known extracellular matrix molecule released into the blood, and increased levels of circulating COL10A1 protein has been suggested as a diagnostic marker of breast cancer^35^. MYOC has been previously reported as a topranked downregulated gene in breast cancer^36^. MMP11 overexpression in early stages is necessary for cancer progression via inhibition of apoptosis, and promotion of invasion and metastasis^37^. Overexpression of LYVE1 has been suggested as a reliable marker of lymphatic metastasis in breast cancer patients^38^. HSD17B13 is involved in estrogen biosynthesis^39^, and its tumor suppressor role in hepatocellular carcinoma has been documented^40^, suggesting analogous key roles specific to breast cancer progression.

Due to the substantial heterogeneity in breast cancer, large feature spaces have been necessary for acceptable performance in contemporary classification strategies. Some of these have mandated whole genome sequencing to completely cover the biomarker space of interest. For e.g, Zhao et al identified 817 features and used them to build a model that achieved accuracies of 86.96% and 72.46% in different external validation datasets respectively^41^. Mostavi et al used a feature space of 2090 genes for discriminating cancer vs normal, of which 323 biomarkers were designated for the task of subtyping breast cancer^42^. Convolution-based deep neural networks (CNNs) have been applied to learn from image datasets of mammography, computed tomography (CT), magnetic resonance (MR) and histopathological slides^43-45^. CNNs have been used to extract features from whole-slide tissue-biopsy images, which were subsequently used to train a Support Vector Machine classifier of cancer vs normal, yielding an accuracy of 83.3%^46^. Radiogenomics approaches based on multimodal datasets have also been developed for breast cancer diagnosis^47^. The use of large feature spaces discourages the use of AI-assisted diagnosis in medical decision-making. Very recently Taghizadeh et al have advanced a solution to the ‘cancer’ vs. ‘normal’ problem, proposing a panel of 20 biomarkers for discriminating breast cancer from normal sample^48^. Their study has been validated on an internal test set with a balanced accuracy ∼ 86%, but no external validation has been provided. Furthermore their models have not been made available for wider use. It is notable that there is zero overlap between biomarkers identified in their study and identified herein. It is hoped that our study provides a reliable and replicable remedy to the present situation, with a balanced-accuracy performance > 95% on the external validation.

## Conclusion

In this work, we set out to negotiate the compromise between model complexity and performance, and develop the simplest possible best-performing model of breast cancer classification. The designed computational pipeline yielded a novel non-redundant hypothesis space of nine biomarkers, which was transformed into a space defined by an optimal number of principal components. A k-means clustering model trained in this transformed space was able to discriminate cancer from normal samples with a high balanced accuracy of 99.5% and 95.5% on the internal and external validation datasets, respectively. At the same time, we note that the model had limited recall (< 92%) on the external validation dataset. The model could be further improved by efforts to predict the subtype of breast cancer as well as its progression to advanced stages or metastasis. The present model has been deployed as a web-service at https://apalania.shinyapps.io/brcadx/ for non-commercial use. The ideas used in our study could be useful in developing elegant, interpretable AI-assisted diagnostic models for many other cancers and disease conditions, fostering effective aid to medical decision-making.

## Data Availability

All data produced in the present work are contained in the manuscript. Software-based diagnostic aid is available at: https://apalania.shinyapps.io/brcadx/

## Acknowledgements

We would like to thank the School of Chemical and Biotechnology and CeNTAB, SASTRA Deemed University, for infrastructure and computing support. This work was partially supported by DST-SERB grant EMR/2017/000470, Government of India.

